# Viral and Symptom Rebound in Untreated COVID-19 Infection

**DOI:** 10.1101/2022.08.01.22278278

**Authors:** Rinki Deo, Manish C. Choudhary, Carlee Moser, Justin Ritz, Eric S. Daar, David A. Wohl, Alexander L. Greninger, Joseph J. Eron, Judith S. Currier, Michael D. Hughes, Davey M. Smith, Kara W. Chew, Jonathan Z. Li, the ACTIV-2/A5401 Study Team

**Affiliations:** Brigham and Women’s Hospital, Harvard Medical School, Boston, MA; Harvard T.H. Chan School of Public Health, Boston, MA; Lundquist Institute at Harbor-University of California, Los Angeles Medical Center, Torrance, CA; University of North Carolina, Chapel Hill, NC; University of Washington Medical Center, Seattle, WA; David Geffen School of Medicine at University of California, Los Angeles, Los Angeles, CA; University of California, San Diego, San Diego, CA

**Keywords:** COVID-19, viral rebound, symptom rebound, Paxlovid

## Abstract

**Background:** There are reports of viral RNA and symptom rebound in people with COVID-19 treated with nirmatrelvir/ritonavir. Since the natural course of viral and symptom trajectories of COVID-19 has not been well described, we evaluated the incidence of viral and symptom rebound in untreated outpatients with mild-moderate COVID-19.

**Methods:** The study population included 568 participants enrolled in the ACTIV-2/A5401 platform trial who received placebo. Anterior nasal swabs were collected for SARS-CoV-2 RNA testing on days 0-14, 21 and 28. Participants recorded the severity of 13 targeted symptoms daily from day 0 to 28. Viral rebound was defined as ≥0.5 log_10_ viral RNA copies/mL increase and symptom rebound was defined as a 4-point total symptom score increase from baseline. Baseline was defined as study day 4 (primary analysis) or 8 days from symptom onset (secondary analysis).

**Findings:** In both the primary and secondary analyses, 12% of participants had viral rebound. Viral rebounders were older than non-rebounders (median 54 vs 47 years, P=0.04). Symptom rebound occurred in 27% of participants after initial symptom improvement and in 10% of participants after initial symptom resolution. The combination of high-level viral rebound to ≥5.0 log_10_ RNA copies/mL and symptom rebound after initial improvement was observed in 1-2% of participants.

**Interpretation:** Viral RNA rebound or symptom relapse in the absence of antiviral treatment is common, but the combination of high-level viral and symptom rebound is rare.

**Funding:** This study was supported by the National Institute of Allergy and Infectious Diseases; ACTIV-2/A5401 ClinicalTrials.gov number NCT04518410.

## INTRODUCTION

Nirmatrelvir-ritonavir (Paxlovid) is a recommended treatment of choice for outpatients with mild-moderate COVID-19 and risk factors for severe disease^1^. With wide-spread use of nirmatrelvir-ritonavir, there have been case reports of individuals experiencing worsening symptoms^2^ and/or virologic rebound^3,4^ after treatment completion (known as post-Paxlovid rebound). However, many questions remain unanswered questions regarding this phenomenon. For example, the natural recovery from COVID-19 does not always progress in a linear fashion and clinical relapses can occur in the absence of antiviral treatment^5,6^. Case reports and case series may be subject to reporting bias and the incidence of viral and symptom rebound is difficult to determine as the denominator is challenging to estimate. Understanding the frequencies of viral and symptom rebound in the absence of treatment is required to fully define the possible role that antiviral therapy may play in these observations. To date, much of the reported literature is observational and has been limited due to the lack of systematically collected samples and data in a rigorous clinical trial setting. Even in the analysis of clinical trials like EPIC-HR, the phase 3 study of nirmatrelvir-ritonavir in outpatients with mild-moderate COVID-19, the frequency of viral rebound is likely underestimated as viral RNA quantification was only performed at two follow-up time points after the completion of the nirmatrelvir-ritonavir or placebo course and symptom rebound was not described.

In this study, we evaluated the incidence of viral and symptom rebound in untreated outpatients with mild-moderate COVID-19 through an analysis of data from participants who received a placebo in the ACTIV-2/AIDS Clinical Trials Group A501 (A5401) multicenter phase 2/3 platform randomized trial. A strength of this study was that participants had daily anterior nasal (AN) sampling for the first two weeks (in the phase 2 studies) for quantitative viral load testing and daily symptom diaries for the first 29 days (in the phase 2 and 3 studies). This intensive sampling in a rigorous randomized, placebo-controlled trial framework allowed an in-depth assessment of the frequencies of viral and symptom rebound after initial improvement for untreated individuals.

## METHODS

### Overview of study participants

Adults (≥18 years) were enrolled in the ACTIV-2/A5401 platform trial for outpatients with mild-moderate COVID-19 (NCT04518410). Viral rebound analysis was restricted to participants who enrolled in the placebo arms of the following ACTIV-2/A5401 phase 2 studies: bamlanivimab 7000 mg (N=46), bamlanivimab 700mg (N=112), and amubarvimab plus romlusevimab (N=109) monoclonal antibodies. Daily self-reported symptoms were collected for the first 28 days. For the symptom rebound analysis, an additional 301 participants were included from the placebo arm of the phase 3 trial of the amubarvimab plus romlusevimab monoclonal antibodies. The bamlanivimab studies enrolled participants who were at standard and higher risk for progression to severe COVID-19 while the amubarvimab plus romlusevimab studies enrolled only high-risk participants. All participants in the phase 2 studies were enrolled in the US while participants in the amubarvimab plus romlusevimab phase 3 evaluation were enrolled in the US, Argentina, Mexico, South Africa and Brazil.

### Definition of “baseline” time point for both viral and symptom analysis

For both the viral and symptom rebound calculations, we have defined a simulated post-nirmatrelvir/ritonavir baseline time point in our analyses that is comparable to the baseline time point in the analysis of viral rebound from the phase 3 trial of nirmatrelvir-ritonavir (EPIC-HR)^7^. For the primary analysis, we restricted to participants with #5 days of symptoms at the time of study enrollment and then designated the 5^th^ day of the study (study day 4) as the baseline time point (Supplementary Table 1). We also considered an alternative definition of baseline in the secondary analyses of viral and symptom rebound where baseline was defined as the 8^th^ day of symptoms (days since symptom onset [DSSO] 8) (Supplementary Table 1). In contrast to the primary analysis, the secondary analysis does not restrict to participants enrolled with ≤5 days of symptom onset but includes all participants with a study visit at DSSO 8. DSSO 8 also simulates the post-treatment timing for participants of the EPIC-HR study, who had a median of 3 days of symptoms before starting their 5 days of nirmatrelvir-ritonavir or placebo (i.e., the EPIC-HR participants had a median of 8 days of symptoms at the time of completion of nirmatrelvir-ritonavir).

### SARS-CoV-2 viral rebound analysis

Daily anterior nasal (AN) swabs were obtained from study entry (day 0) through study day 14 and at day 28. For bamlanivimab participants, in addition to the above, an additional sample at day 21 were also collected for viral load testing. SARS-CoV-2 RNA levels were quantified from AN swab sample using the Abbott m2000 system and a Laboratory Developed Test (LDT) as previously described^8-10^. Participants were included in the analysis if SARS-CoV-2 RNA levels were available from at least 3 time points (the median [Q1, Q3] number of viral RNA measurements per participant was 16 [14, 16]). Viral rebound was defined as ≥0.5 log_10_ increase in AN SARS-CoV-2 RNA at a follow-up time point relative to baseline, with the follow-up RNA level meeting a certain threshold (Supplementary Figure 1). We considered the frequency of viral rebound at a minimum viral RNA rebound level or at least 3.0 and 5.0 log_10_ RNA copies/mL. The 3.0 log_10_ threshold is similar to the one used in the analysis of viral rebound in EPIC-HR while the 5 log_10_ copies/mL threshold was chosen as our previous studies have demonstrated a high rate of SARS-CoV-2 culture positivity at ≥ 5.0 log_10_ SARS-CoV-2 RNA copies/mL^11^, which may have transmission implications.

### Symptom score rebound analysis

Total symptom scores were calculated on each day as the sum of scores for 13-targeted symptoms, based on a daily self-collected symptom diary from day 0 to 28. The targeted symptoms included feverishness, cough, shortness of breath or difficulty breathing, sore throat, body pain or muscle pain or aches, fatigue, headache, chills, nasal obstruction or congestion, nasal discharge, nausea, vomiting, and diarrhea. Each symptom was self-assessed and scored daily by the participant as absent (assigned 0 points), mild (1), moderate (2), or severe (3). The total symptoms scores for each day were calculated by summing the individual scores for all 13 symptoms. Missing responses for individual symptoms were ignored in calculation of the total symptom score.

We evaluated symptom rebound in two ways: total symptom score increase of ≥4 points after initial improvement and total symptom rebound increase of ≥4 points after symptom resolution (defined as symptom score reaching ≤2 points). To identify participants with symptom rebound after improvement, the following steps were taken: 1) the maximum total symptom score after baseline (“maximum score”) was identified, 2) the minimum total symptom score between baseline and the maximum score (“minimum score”) was identified, 3) symptom improvement was determined by the participant having a symptom score higher than the minimum score between baseline and the minimum score, and 4) the magnitude of symptom rebound was calculated as the difference between minimum and maximum scores (Supplementary Figure 2A, 2B). Participants were excluded if hospitalization occurred on or before the baseline time point or there was no evidence of symptom improvement prior to hospitalization.

To identify cases of symptom rebound after resolution, the following steps were taken: 1) the first time point with a total symptom score ≤2 after the baseline time point (“first symptom resolution time point”) was identified, 2) it was confirmed that prior to the first resolution time point, there was a time point with a higher symptom score, 3) the maximum symptom score after first symptom resolution time point (“maximum score”) was identified, 4) the minimum symptom score between the first symptom resolution time point and the maximum score time point (“minimum score”) was identified, and 5) the magnitude of symptom rebound was calculated as the difference between the minimum and maximum scores (Supplementary Figure 2C, 2D). Hospitalized participants were included if the hospitalization occurred after baseline and there was a pre-hospitalization symptom score ≤2. Since the onset of high-level SARS-CoV-2 RNA shedding and symptoms are frequently offset during acute COVID-19, the calculation of the frequency both viral and symptom rebound included individuals meeting viral and symptom rebound definitions at any time point after baseline.

### Statistical analysis

SARS-CoV-2 RNA level below the limit of detection (LoD) were imputed as 0.7 log_10_ copies/ml, while lower limit of quantification (LLoQ) was imputed as 1.7 log_10_ copies/ml. Continuous variables are presented as medians with inter-quartile range, while categorical variables are expressed as frequencies or percentages. Statistical analysis was performed using Mann Whitney U tests for continuous variables and Fisher’s exact tests for discrete variables. All statistical analyses were performed in GraphPad Prism (Version 9.1.1).

## RESULTS

In the primary viral and symptom rebound analysis, we used the 5^th^ day since study enrollment (study day 4) as the baseline time point as it simulates the end of a 5-day treatment course with nirmatrelvir/ritonavir (Supplemental Figure 1). Eleven (12%) participants were found to have viral rebound of ≥0.5 log_10_ RNA copies/ml rebound at a post-study day 4 time point, with a minimum SARS-CoV-2 RNA rebound of ≥3.0 log_10_ copies/mL (Figure 1a). The majority of viral rebound ≥3.0 log_10_ occurred within the first 5 days after baseline (73%) and lasted for 1 day (91%). Although non-significant, individuals with viral RNA rebounders had higher baseline AN viral RNA levels and had detectable median AN SARS-CoV-2 RNA level for longer duration (Figure 1B). Viral RNA rebounders were found to be older than non-rebounders (median 54 vs 47 years, P=0.04, Table 1). There were no significant differences in sex, race, days since symptom onset to enrollment, or symptom scores at enrollment between those with and without viral rebound. We also evaluated the frequency of high-level nasal RNA rebound with a minimum rebound threshold of 5.0 log_10_ and we found that 5.3% (n=5) participants met this definition of viral rebound (Figure 1a). We also performed a supplementary analysis using the alternative definition of the baseline: DSSO 8 (Supplementary table 1). A total of 204 participants were included in this supplementary analysis. The results were similar to the primary analysis, demonstrating 12% (n=24) and 6.9% (n=14) participants had viral rebound rates based on minimum thresholds of ≥3.0 and 5.0 log_10_ RNA copies/mL, respectively (Figure 1A).

**Figure 1:**
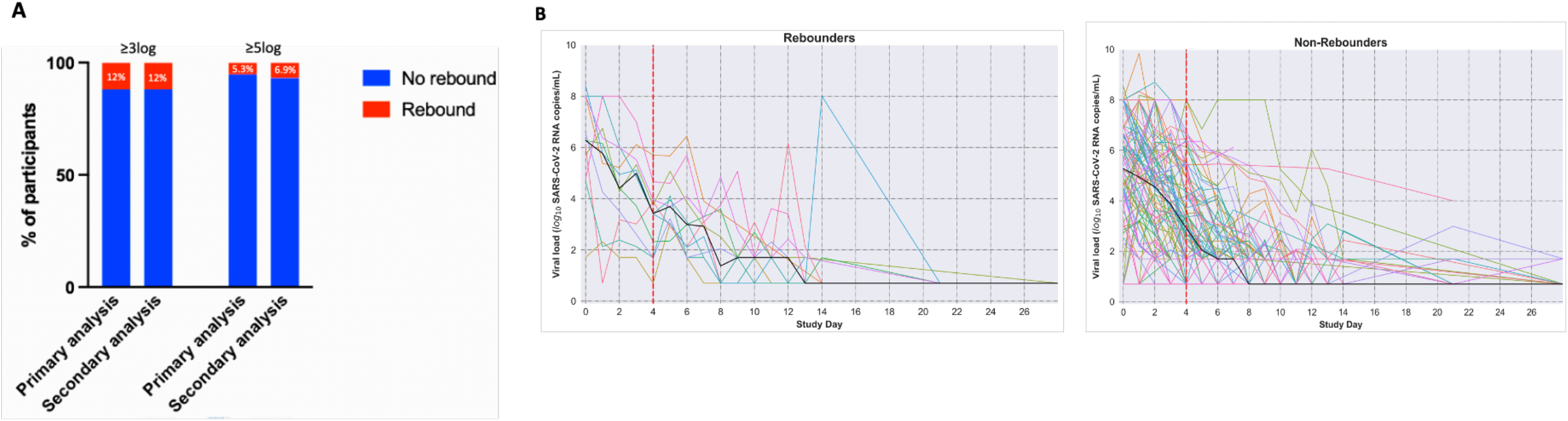
Description of anterior nasal (AN) SARS-CoV-2 RNA rebound. (A) Bar graph shows percentage of participants having ≥0.5 log_10_ AN SARS-CoV-2 RNA rebound at a follow-up time point relative to baseline using the primary (study day 4) and secondary analysis (8 days from symptom onset). The frequencies of viral rebound were assessed with a minimum rebound viral load of either ≥3.0 or ≥5.0 log_10_ RNA copies/mL. (B) The left and right graphs show log_10_ AN SARS-CoV-2 RNA in copies/ml by study day in rebounders and non-rebounders respectively using primary definition of baseline i.e, study day 4 and rebound viral load value ≥3 log AN SARS-CoV-2 RNA copies/ml. Median AN SARS-CoV-2 RNA copies/ml for each day is shown with thick black line. Y-axis shows log_10_ AN SARS-CoV-2 RNA in copies/ml while x-axis denotes study day.

**Table 1:** Demographic characteristics of participants categorized as rebounders and non-rebounders using viral load and total symptom score criteria. Viral rebounders were defined as individuals with ≥0.5 log_10_ SARS-CoV-2 RNA copies/mL increase from study day 4. Symptom rebound was defined as an increase of ≥4 points on the total symptom score from study day 4. Statistical analysis was performed using Mann Whitney U tests for continuous variables and Fisher’s exact tests for discrete variables. p-values which are significant are shown as bold. AN: anterior nasal.

| Nasal viral rebound analysis (N=95) |  |  |  | Symptom rebound analysis (N=247) |  |  |
| --- | --- | --- | --- | --- | --- | --- |
| Characteristic | Rebounders<br>(N = 11) | Non-rebounders<br>(N = 84) | P-Value | Rebounders<br>(N = 66) | Non-rebounders<br>(N = 181) | P-Value |
| Age, median years<br>[Q1,Q3] | 54 [50, 64] | 47 [36, 55] | **0.04** | 52 [42, 61] | 48 [38, 57] | 0.08 |
| Female sex, % | 73 | 51 | 0.21 | 62 | 46 | **0.03** |
| Race/Ethnicity, % |  |  | 0.59 |  |  | 0.9 |
| White | 100 | 90 |  | 82 | 81 |  |
| Black | 0 | 5 |  | 8 | 13 |  |
| Asian | 0 | 4 |  | 5 | 3 |  |
| Other | 0 | 1 |  | 5 | 3 |  |
| Baseline AN VL, median<br>log<sub>10</sub> SARS-CoV-2<br>copies/mL [Q1, Q3] | 6.3 [4.8, 8.0] | 5.3 [3.2, 6.9] | 0.10 | 6.5 [4.6, 7.7] | 4.7 [2.6, 6.2] | **0.0006** |
| Days from symptom<br>onset to enrollment,<br>median [Q1, Q3] | 3 [2, 4] | 4 [3, 4] | 0.27 | 4 [3, 4] | 4 [3, 4] | 0.72 |
| Symptom Score at<br>enrollment (Study Day 0) | 9 [6, 13] | 9 [7, 12] | 0.78 | 13 [10, 19] | 9 [6, 13] | **<0.0001** |
| Symptom Score at<br>baseline (Study Day 4) | 4 [2, 7] | 5 [3, 8] | 0.26 | 7.5 [4, 13] | 4 [2, 7] | **<0.0001** |

The median [Q1, Q3] symptom score at study enrollment was 10 points [6, 15]. Using both definitions of baseline, we assessed the frequency of symptom rebound (≥4 point increase in total symptom score) after initial improvement (Supplementary figure 2A, 2B). In the primary analysis population, which includes 247 participants receiving placebo, we found symptom rebound after initial improvement occurred in 27% (n=66) of participants (Figure 2A). Individuals with symptom rebound were more likely to be female, had higher baseline AN viral RNA levels and higher symptom scores at both study enrollment and baseline time points (Table 1). There were no significant differences in race/ethnicity or days from symptom onset to enrollment. Evaluating the frequency of symptom rebound after initial symptom resolution, we found that 10% (n=22) participants in the primary analysis met the definition of symptom rebound after resolution. The results were consistent with supplementary analysis using the alternative definition of baseline (DSSO 8). This analysis included 428 participants, of whom 25% (n=106) had symptom rebound after improvement and 16% (n=58) had symptom rebound after initial resolution (Figure 2B).

**Figure 2:**
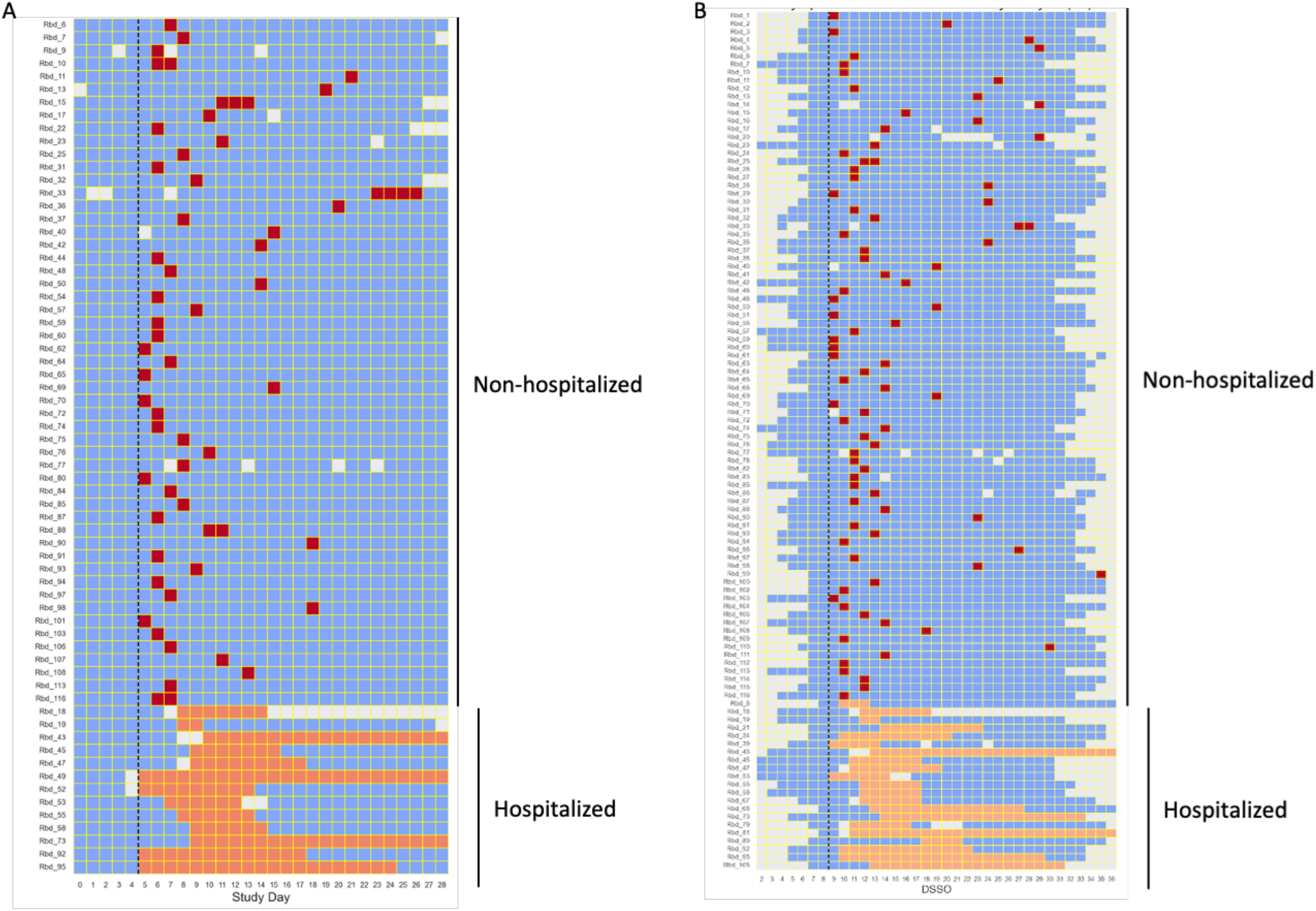
Heat map of symptom score rebound. The heat map shows participants with hospitalization or symptom rebound (≥4-point increase from baseline in symptom score) after demonstrating initial symptom improvement using (A) study day 4 as baseline, (B) day 8 after symptom onset as baseline. Baseline time point is shown as dotted black line. Individual participants are shown in rows while study day is shown in columns. Red squares represent days meeting symptom rebound criteria and orange squares represent days of hospitalization. Blue squares denote days that did not meet symptom rebound criteria.

Finally, we assessed the frequency of individuals meeting both the viral and symptom rebound criteria. This analysis was restricted to participants with both daily nasal SARS-CoV-2 RNA and symptom score measurements (n=93 and n=173 for primary secondary analysis populations, respectively). While symptom rebound was commonly seen, the combination of both viral and symptom rebound was rare (Table 2). For example, high-level viral rebound ≥5.0 log_10_ RNA copies/mL along with symptom rebound after improvement was detected in 2.2% (n=2) and 1.2% (n=2) of participants using either the primary or supplementary baseline definitions, respectively. No participants had both viral rebound ≥5.0 log_10_ RNA copies/mL and symptom rebound after initial symptom resolution.

**Table 2:**
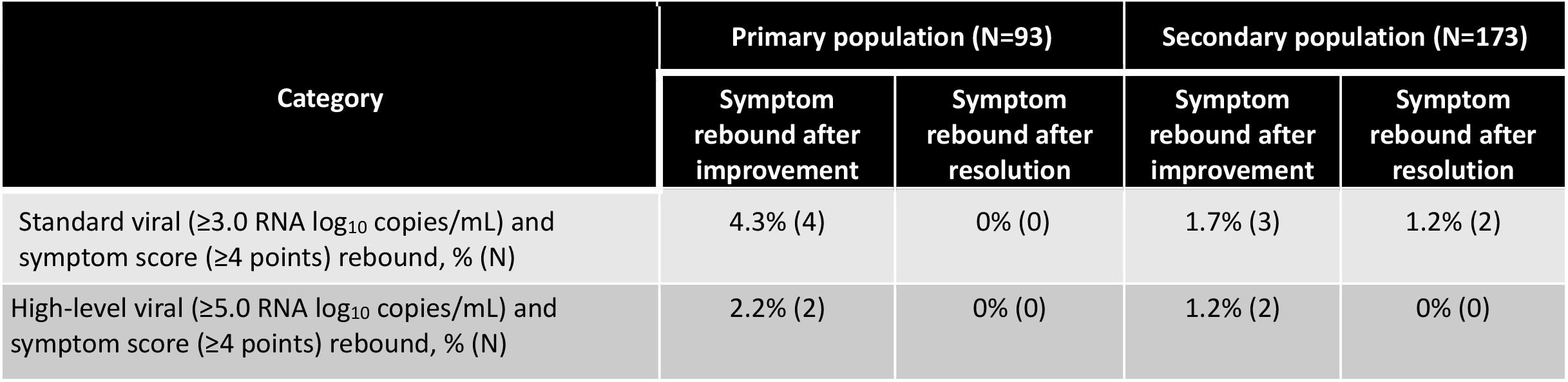
Frequency of participants meeting both viral and symptom rebound criteria using either the primary (study day 4 as baseline) or secondary (8 days since symptom onset as baseline) analysis definitions. Symptom rebound was assessed following either symptom resolution or improvement.

| Category | Primary population (N=93) |  | Secondary population (N=173) |  |
| --- | --- | --- | --- | --- |
|  | Symptom rebound after improvement | Symptom rebound after resolution | Symptom rebound after improvement | Symptom rebound after resolution |
| Standard viral ($\geq 3.0$ RNA $\log_{10}$ copies/mL) and symptom score ($\geq 4$ points) rebound, % (N) | 4.3% (4) | 0% (0) | 1.7% (3) | 1.2% (2) |
| High-level viral ($\geq 5.0$ RNA $\log_{10}$ copies/mL) and symptom score ($\geq 4$ points) rebound, % (N) | 2.2% (2) | 0% (0) | 1.2% (2) | 0% (0) |

## DISCUSSION

In this study of ACTIV-2/A5401 randomized controlled trial participants who received placebo and had daily nasal sampling for SARS-CoV-2 RNA and symptom assessment, we found that viral or symptom rebound after initial improvement was relatively common, with 1 in 8 individuals experiencing a viral rebound and 1 in 4 participants experiencing symptom relapse. However, the duration of viral rebound was short, lasting 1 day for the vast majority of individuals, and the frequency of individuals meeting both high-level viral (≥5.0 log_10_ RNA copies/mL) and symptom rebound criteria was uncommon, occurring in ≤2% of study participants receiving placebo in the trial.

With the anecdotal reports of clinical relapse after nirmatrelvir-ritonavir treatment, it is important to understand the natural history of COVID-19 and underlying rates of viral and symptom rebound. In the analysis of the EPIC-HR phase 3 outpatient study of nirmatrelvir-ritonavir for mild-moderate COVID-19, a 0.5 log_10_ or greater increase in nasal SARS-CoV-2 RNA levels from post-treatment levels was detected in approximately 4% of participants receiving placebo and 7% of participants receiving nirmatrelvir-ritonavir^7^. However, viral RNA levels were only quantified at two follow-up time points (5 and 9 days after the end of nirmatrelvir-ritonavir/placebo) and this may explain why we found a much higher rate of viral rebound with intensive daily sampling. With daily nasal viral quantification at up to 16 follow-up time points, we showed that a 0.5 log_10_ or greater increase in SARS-CoV-2 RNA occurred in 12% of untreated participants using two different definitions of baseline that were chosen to be analogous to baseline in the EPIC-HR study.

Published cases of clinical relapse after nirmatrelvir-ritonavir have described both symptom rebound and the recurrence of culture positive virus^3,4^. However, the EPIC-HR study did not evaluate rates of high-level viral rebound that might be associated with culture positive virus and did not report rates of symptom rebound. In this study of untreated individuals, we found that symptom rebound after initial improvement was common, occurring in approximately 25% of participants and that symptom rebound after resolution was experienced by 10-16%. We also identified characteristics associated with the occurrence of symptom rebound, including female sex and higher levels of nasal SARS-CoV-2 RNA shedding and higher symptom scores at study enrollment.

We had previously demonstrated that culture positive virus is commonly detected when SARS-CoV-2 RNA levels are ≥5.0 log_10_ copies/mL^11^. In an analysis of participants with both high-level SARS-CoV-2 RNA rebound (≥5.0 log_10_ copies/mL) and symptom rebound, only 1-2% had evidence of symptom rebound after initial symptom improvement; no participants had both high level viral rebound and symptom rebound after resolution. Together, these results show that while waxing and waning symptom course may be commonly reported, symptom relapse with high-level viral load rebound is rare.

There are several potential etiologies for the relapsing symptoms described here during acute SARS-CoV-2 acute infection. One possibility is viral dissemination into different anatomic compartments over time that could cause an evolving series of symptoms^13,14^. In the setting of high-levels of community COVID-19 infection, infection with two separate SARS-CoV-2 variants has been described, although this is still thought to be a relatively rare occurrence^15^. In addition, co-infection with another respiratory virus is a possibility, along with symptom rebound from a non-infectious etiology. Given its high frequency, symptom rebound during acute COVID-19 is likely to be multifactorial.

This study has some limitations. The results could be affected by the underlying study population as the ACTIV-2/A5401 study enrolled a largely unvaccinated population infected with pre-Omicron variants, including a subset of individuals without risk factors for severe COVID-19. Of note, recently published studies have reported that neither vaccination nor Omicron variant substantially alters viral decay kinetics^11,16^. As the ACTIV-2/A5401 study did not enroll participants receiving nirmatrelvir-ritonavir, we are unable to define rates of post-treatment viral or symptom rebound. For individuals experiencing symptom relapse after completion of nirmatrelvir-ritonavir, a maturing immune response reacting to the sudden re-appearance of viral antigen could be an important contributory factor^3,12^. Our results highlight, though, the importance of accounting for underlying rates of symptom relapse in the absence of antiviral therapy when evaluating the effects of treatment with nirmatrelvir-ritonavir or other antiviral agents.

In summary, we observed that viral RNA rebound and symptom score rebound is relatively common in participants who are not treated with any antiviral agents. Viral rebounders were older, while symptom rebounders were more likely to be female and have higher AN viral RNA levels and symptom scores at study enrollment. However, co-occurrence of both high-level viral and symptom rebound was rare. These results provide insight into the natural trajectory of viral rebound and symptom relapses during COVID-19, which is critical in the interpretation of studies reporting biphasic disease courses after nirmatrelvir/ritonavir and other antiviral treatments.

## Data Availability

Data are available under restricted access due to ethical restrictions. Access can be requested by submitting a data request at https://submit.mis.s-3.net/ and will require the written agreement of the AIDS Clinical Trials Group (ACTG) and the manufacturer of the investigational product. Requests will be addressed as per ACTG standard operating procedures. Completion of an ACTG Data Use Agreement may be required. All analyses were performed using code available in standard software packages. No new code was developed for this manuscript.

## Contributors

R.D, M.C.C., M.D.H., D.M.S., K.W.C., J.Z.L. conceptualized and performed the study, R.D., M.C.C., C.M., J.R., performed analysis; A.L.G. performed viral load testing. E.S.D., D.A.W., J.J.E., J.S.C., M.D.H., D.M.S., provided critical inputs; R.D., M.C.C, J.Z.L., performed the statistical analysis, all authors contributed to manuscript writing and editing.

## Acknowledgements

This work was supported by the National Institute of Allergy and Infectious Diseases of the National Institutes of Health under Award Number UM1 AI106701, UM1AI068634 (MDH), UM1AI068636 (JSC) and UM1AI106701 (GA). The content is solely the responsibility of the authors and does not necessarily represent the official views of the National Institutes of Health. The authors thank the study participants, site staff, site investigators, and the entire ACTIV-2/A5401 study team; the AIDS Clinical Trials Group, including Lara Hosey, Jhoanna Roa, and Nilam Patel; [the UW Virology Specialty Laboratory staff, including Emily Degli-Angeli, Erin Goecker, Glenda Daza, Socorro Harb, and Joan Dragavon; the ACTG Laboratory Center, including Grace Aldrovandi and William Murtaugh; Frontier Science, including Marlene Cooper, Howard Gutzman, Kevin Knowles, and Rachel Bowman; the Harvard Center for Biostatistics in AIDS Research (CBAR) and ACTG Statistical and Data Analysis Center (SDAC); the National Institute of Allergy and Infectious Diseases (NIAID) / Division of AIDS (DAIDS); Bill Erhardt; the Foundation for the National Institutes of Health and the Accelerating COVID-19 Therapeutic Interventions and Vaccines (ACTIV) partnership, including Stacey Adams; and the PPD clinical research business of Thermo Fisher Scientific. The authors also thank the members of the ACTIV-2/A5401 data and safety monitoring board — Graeme A. Meintjes, PhD, MBChB (Chair), Barbara E. Murray, MD, Stuart Campbell Ray, MD, Valeria Cavalcanti Rolla, MD, PhD, Haroon Saloojee, MB.BCh, FCPaed, MSc, Anastasios A. Tsiatis, PhD, Paul A. Volberding, MD, Jonathan Kimmelman, PhD, David Glidden, PhD, and Sally Hunsberger, PhD (Executive Secretary).

---

**Supplementary Table 1:** Table showing definition of study population and baseline time point used using primary and secondary analysis.

| Analysis | Study Population | Baseline time point |
| --- | --- | --- |
| Primary | Participants with $\leq 5$ days of symptoms at study entry and data available at study day 4 | Study day 4 |
| Secondary | Data available at day 8 of symptom onset | Day 8 after symptom onset |

**Supplementary Figure 1:**
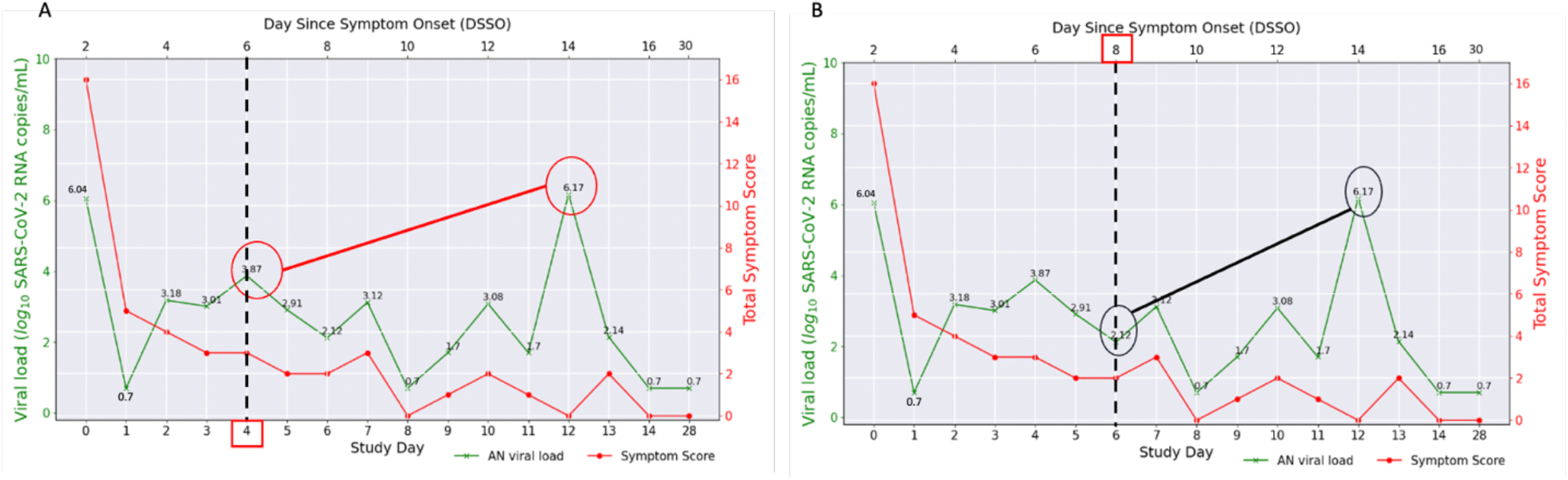
Example of assessments of viral rebound. An example of viral load rebound case (A) using study day 4 as baseline or (B) using days since symptom onset 8 as baseline. Left y-axis denotes AN SARS-CoV-2 RNA copies/ml (green line graph) and right y-axis denotes total symptom score (red line graph) while bottom x-axis shows study day and top x-axis shows days since symptom onset. Encircled values in (A) and (B) shows baseline viral load value and highest viral load at follow-up time-point. Baseline time point in figure A and B are represented by red square box.

**Supplementary Figure 2:**
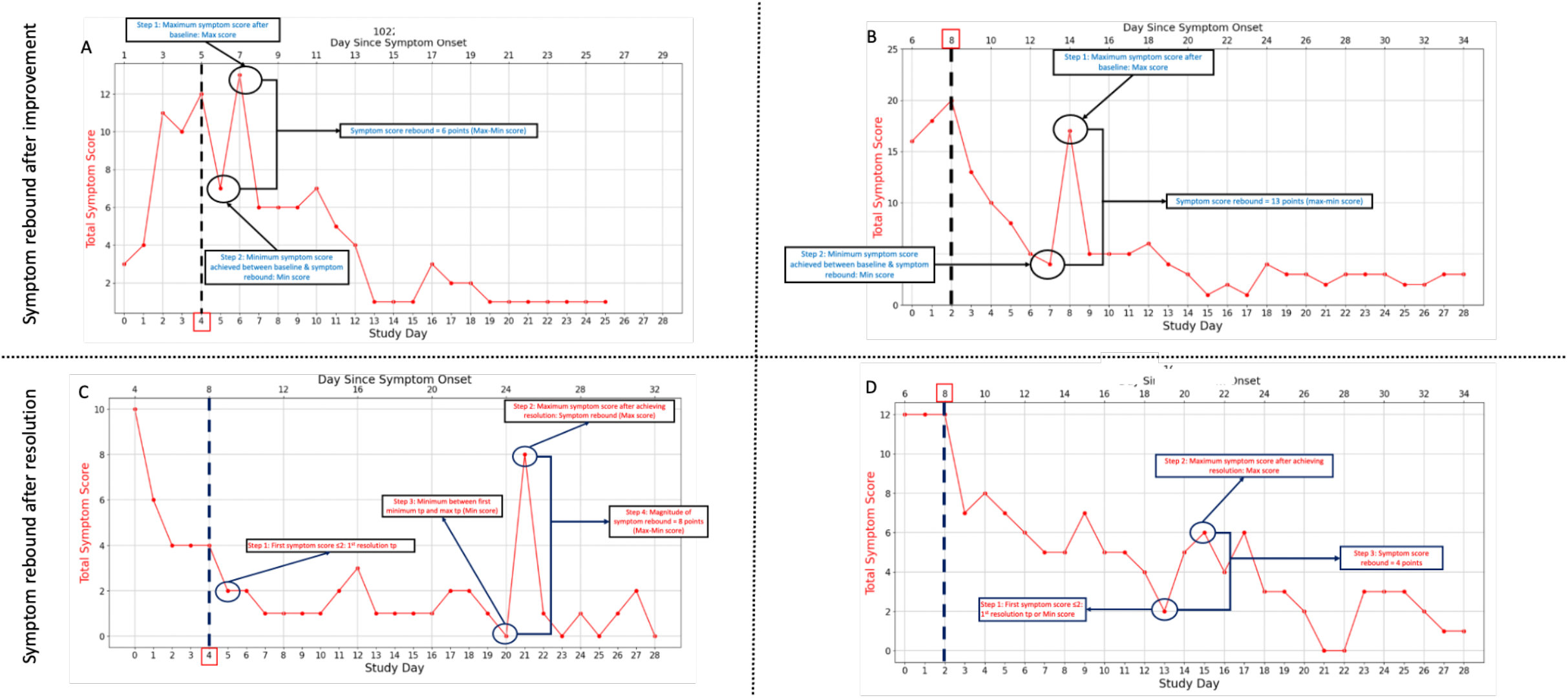
Example of symptom rebounders by primary and secondary definition of baseline. The upper panels (A, B) show an example of symptom rebound after initial symptom improvement by the two different definitions of baseline, (A) baseline defined as study day 4 (primary definition) and (B) baseline defined as days since symptom onset 8 (secondary definition), while the lower panels C and D show an example of symptom rebound after initial symptom resolution with study day 4 and days since symptom onset 8 as baseline, respectively. Y-axis denotes total symptom score while lower x-axis shows study day and upper x-axis shows days since symptom onset. Baseline is shown by thick dashed vertical line while encircled points are the symptom score values chosen to calculate symptom rebound after baseline.

## References

1. Hammond J, Leister-Tebbe H, Gardner A, et al. Oral Nirmatrelvir for High-Risk, Nonhospitalized Adults with Covid-19. N Engl J Med 2022; 386(15): 1397–408.

2. Ranganath N, O’Horo JC, Challener DW, et al. Rebound Phenomenon after Nirmatrelvir/Ritonavir Treatment of Coronavirus Disease-2019 in High-Risk Persons. Clin Infect Dis 2022.

3. Carlin AF, Clark AE, Chaillon A, et al. Virologic and Immunologic Characterization of COVID-19 Recrudescence after Nirmatrelvir/Ritonavir Treatment. Clin Infect Dis 2022.

4. Boucau J, Uddin R, Marino C, et al. Virologic characterization of symptom rebound following nirmatrelvir-ritonavir treatment for COVID-19. medRxiv 2022: 2022.05.24.22275326.

5. Gousseff M, Penot P, Gallay L, et al. Clinical recurrences of COVID-19 symptoms after recovery: Viral relapse, reinfection or inflammatory rebound? J Infect 2020; 81(5): 816–46.

6. Buskermolen M, Te Paske K, van Beek J, et al. Relapse in the first 8 weeks after onset of COVID-19 disease in outpatients: Viral reactivation or inflammatory rebound? J Infect 2021; 83(2): e6–e8.

7. Soares H, Baniecki M, Cardin R, et al. Viral Load Rebound in Placebo and Nirmatrelvir-Ritonavir Treated COVID-19 Patients is not Associated with Recurrence of Severe Disease or Mutations. Research Square 2022.

8. Degli-Angeli E, Dragavon J, Huang ML, et al. Validation and verification of the Abbott RealTime SARS-CoV-2 assay analytical and clinical performance. J Clin Virol 2020; 129: 104474.

9. Choudhary MC, Chew KW, Deo R, et al. Emergence of SARS-CoV-2 Resistance with Monoclonal Antibody Therapy. medRxiv 2021.

10. Boucau J, Chew KW, Choudhary MC, et al. Monoclonal antibody treatment drives rapid culture conversion in SARS-CoV-2 infection. Cell Rep Med 2022: 100678.

11. Boucau J, Marino C, Regan J, et al. Duration of viable virus shedding in SARS-CoV-2 omicron variant infection. medRxiv 2022.

12. Epling BP, Rocco JM, Boswell KL, et al. COVID-19 redux: clinical, virologic, and immunologic evaluation of clinical rebound after nirmatrelvir/ritonavir. medRxiv 2022.

13. Fajnzylber J, Regan J, Coxen K, et al. SARS-CoV-2 viral load is associated with increased disease severity and mortality. Nat Commun 2020; 11(1): 5493.

14. Regan J, Flynn JP, Rosenthal A, et al. Viral Load Kinetics of Severe Acute Respiratory Syndrome Coronavirus 2 in Hospitalized Individuals With Coronavirus Disease 2019. Open Forum Infect Dis 2021; 8(8): ofab153.

15. Lacek KA, Rambo-Martin BL, Batra D, et al. SARS-CoV-2 Delta-Omicron Recombinant Viruses, United States. Emerg Infect Dis 2022; 28(7): 1442–5.

16. Kandel CE, Banete A, Taylor M, et al. Similar duration of viral shedding of the severe acute respiratory coronavirus virus 2 (SARS-CoV-2) delta variant between vaccinated and incompletely vaccinated individuals. Infect Control Hosp Epidemiol 2022: 1–3.

